# Drivers of COVID-19 policy stringency in 175 countries and territories: COVID-19 cases and deaths, gross domestic products per capita, and health expenditures

**DOI:** 10.1101/2022.07.05.22277269

**Authors:** Mohamed F. Jalloh, Zangin Zeebari, Sophia A. Nur, Dimitri Prybylski, Aasli A. Nur, Avi J. Hakim, Maike Winters, Laura C. Steinhardt, Wangeci Gatei, Saad B. Omer, Noel T. Brewer, Helena Nordenstedt

**Affiliations:** Center for Global Health, U.S. Centers for Disease Control and Prevention, Atlanta, Georgia, USA; Department of Global Public Health, Karolinska Institute, Stockholm, Sweden; Jönköping International Business School, Jönköping University, Jönköping, Sweden; Department of Sociology, University of Washington, Seattle, Washington, USA; Institute for Global Health, Yale University, New Haven, Connecticut, USA; Department of Health Behavior, Gillings School of Global Public Health, University of North Carolina, Chapel Hill, North Carolina, USA; Lineberger Comprehensive Cancer Center, University of North Carolina, Chapel Hill, North Carolina, USA; Department of Internal Medicine and Infectious Diseases, Danderyd University Hospital, Stockholm, Sweden

**Keywords:** COVID-19, stringency, policy response, public health and social measures, non-pharmaceutical interventions

## Abstract

**Objective:** To understand the associations of COVID-19 cases and deaths with policy stringency globally and regionally.

**Methods:** We modeled the marginal effects of new COVID-19 cases and deaths on policy stringency (scored 0–100) in 175 countries and territories, adjusting for gross domestic product (GDP) per capita and health expenditure (% of GDP). Time periods examined were March–August 2020, September 2020– February 2021, and March–August 2021.

**Results:** Policy response to new cases and deaths was faster and more stringent early in the COVID-19 pandemic (March–August 2020) compared to subsequent periods. New deaths were more strongly associated with stringent policies than new cases. In an average week, 1 new death per 100,000 people was associated with a stringency increase of 2.1 units in March–August 2020, 1.3 units in September 2020–February 2021, and 0.7 units in March–August 2021. New deaths in Africa and the Western Pacific were associated with more stringency than in other regions. Higher health expenditure was associated with less stringent policies. GDP per capita did not have consistent patterns of associations with stringency.

**Conclusions:** Our findings demonstrate the need for enhanced mortality surveillance to ensure policy alignment during health emergencies. Countries that invest less of their GDP in health are inclined to enact stringent policies during health emergencies than countries with more significant health expenditure.

## INTRODUCTION

Policy responses during the COVID-19 pandemic have a dynamic relationship with epidemiological outcomes.[1,2] The effects of public health policies in slowing the spread of severe acute respiratory syndrome coronavirus 2 (SARS-COV-2) are demonstrated in multiple studies, and new evidence is emerging.[3-5] Public health policies to contain the spread of SARS-COV-2 have included restricting population movements and gatherings, closing schools and businesses, and requiring masks indoors—also known as non-pharmaceutical interventions (NPI). Numerous platforms track the stringency of policies enacted globally since the early stages of the pandemic.[6] However, the drivers of these policies remain poorly understood.

Mathematical modeling suggests that policies for NPI are critical to contain the COVID-19 pandemic alongside improved access to and uptake of COVID-19 vaccines [7]. The emergence of variants of concern with immune evasion capability, such as Omicron, reinforces the need to continually (re)calibrate public health policies to control the pandemic.[8] These modeling findings are layered on the backdrop of persistent inequities in access to and uptake of COVID-19 vaccines in low-income countries compared to middle- and high-income countries.[9]

Member states report new COVID-19 cases and deaths to the World Health Organization (WHO).[10] The reported epidemiological trends create dynamic perceptions about the intensity of the COVID-19 situation in a country [11]. They may trigger policies that are not aligned with the COVID-19 epidemiology. Moreover, seroprevalence assessments of SARS-CoV-2 have uncovered gross underestimation of the population-level burden of infection when compared to officially reported new cases.[12,13] Variations in testing capacity, testing strategies, and people’s willingness to be tested contribute to the underestimation of SARS-CoV-2 infections.[14] COVID-19-related mortality may also be underreported due to weak mortality surveillance systems, inadequate systems for civil registration of deaths, and differences in how causes of death are classified.[15]

How rapidly and aggressively countries respond to the reported new cases and deaths may also be influenced by socioeconomic conditions and political considerations.[16,17] An empirical understanding of how the reported epidemiological data influences policy stringency can help predict policy responses in the current pandemic and other health emergencies. To this end, we aimed to model the associations of reported COVID-19 new cases and deaths with policy stringency.

## METHODS AND MATERIALS

Our study examined data for 175 countries and territories representing all regions globally. We used fractional regression to model the marginal effects of new cases and deaths on COVID-19 policy stringency, adjusting for the gross domestic product (GDP) per capita and health expenditure. We adjusted for GDP per capita because prior studies have shown a dynamic relationship between GDP and policy stringency.[19] GDP may influence policy stringency tolerable to a government. Protracted ‘lockdown’ policies, for instance, may not be tenable in low GDP countries. We adjusted for health expenditure (as a percent of GDP) because countries with greater health investments may have greater investments in their COVID-19 response.[20] The GDP and the health expenditure do not change remarkably on an annual basis for the same country. We used the latest reported GDP and health expenditure statistics from the United Nations.[21]

### Main outcome

Our main outcome is the stringency of COVID-19 policies as measured daily by the Oxford Tracker [22]; its methods data collection and scoring methods have been described elsewhere.[23] In summary, the stringency index ranges from 0 to 100, with higher scores representing more stringent policies. The score is based on nine component indicators measured on an ordinal scale to assess closing of schools and universities, closing of workplaces, cancelling public events, limits on gatherings, closing of public transport, mandates for shelter-in-place / home-confinement, restrictions on internal movement between cities/regions, restrictions on international travel, and presence of COVID-19 public information campaigns.

### Statistical analysis

We analyzed the data in Stata version 17 SE (StataCorp LLC, College Station, TX) by examining three six-month periods separately for March–August 2020, September 2020–February 2021, and March– August 2021. These three periods were selected a priori, and the start period (March 2020) was the onset of when most countries reported their first COVID-19 case.

We first described each of the nine ordinally measured stringency indicators for each period and combined periods (Table 1).We then merged the daily COVID-19 epidemiological data countries report to the WHO [24] with their policy stringency data.[22] New cases and deaths were calculated to reflect cases and deaths per 100,000 people. The daily merged data for cases, deaths, and stringency were aggregated weekly. We added each country’s/territory’s time-invariant data on GDP per capita [25] and health expenditure to the dataset. Finally, we used fixed-effects fractional regression with a quasi-likelihood method to model the marginal effects [18] of the various covariates on policy stringency.

**Table 1.** Changes in stringency of specific COVID-19 policies—175 countries and territories, March 2020—August 2021.

|  | **Mar 2020–Aug 2020**<br>32,200 daily reports<br>(median=68.52)<br>% | **Sep 2020–Feb 2021**<br>31,675 daily reports<br>(median=56.48)<br>% | **Mar 2021–Aug 2021**<br>34,040 daily reports<br>(median=54.63)<br>% | **Overall**<br>115,325 daily reports<br>(median=55.56)<br>% |
| --- | --- | --- | --- | --- |
| **(C1) Closings of schools and universities** |  |  |  |  |
| 0 – no measures | 11.19 | 10.45 | 20.07 | 22.07 |
| 1 – recommend closing or all schools open with alterations | 9.08 | 34.71 | 34.42 | 23.52 |
| 2 – require closing for some levels or categories | 18.58 | 26.11 | 24.43 | 20.80 |
| 3 – require closing all levels | 61.15 | 28.72 | 20.82 | 33.52 |
| **(C2) Closings of workplaces** |  |  |  |  |
| 0 – no measures | 21.03 | 16.29 | 14.83 | 25.17 |
| 1 – recommend closing or remote work or significant alterations | 13.98 | 24.34 | 27.90 | 20.04 |
| 2 – require closing (or remote work) for some sectors or workers | 46.02 | 48.03 | 45.21 | 41.99 |
| 3 – require closing (or remote work) for all but essential workplaces | 18.97 | 11.30 | 11.82 | 12.72 |
| **(C3) Cancelling of public events** |  |  |  |  |
| 0 – no measures | 13.86 | 11.50 | 10.71 | 20.45 |
| 1 – recommend cancelling | 13.87 | 27.85 | 32.88 | 22.41 |
| 2 – require cancelling | 72.26 | 60.65 | 56.38 | 57.12 |
| **(C4) Limits on gatherings** |  |  |  |  |
| 0 – no restrictions | 18.71 | 12.62 | 11.57 | 22.39 |
| 1 – restrictions on very large gatherings of above 1000 people | 2.68 | 2.66 | 2.87 | 2.46 |
| 2 – restrictions on gatherings between 101 and 1000 people | 10.05 | 11.06 | 6.66 | 8.16 |
| 3 – restrictions on gatherings between 11 and 100 people | 28.28 | 30.61 | 31.72 | 27.26 |
| 4 – restrictions on gatherings of 10 people or less | 40.28 | 43.01 | 47.10 | 39.68 |
| **(C5) Public transport** |  |  |  |  |
| 0 – no measures | 47.09 | 57.71 | 51.52 | 56.72 |
| 1 – recommend closing or significantly reduced | 30.84 | 33.59 | 39.92 | 31.42 |
| 2 – require closing or prohibit most citizens from using it | 22.06 | 8.62 | 8.46 | 11.81 |
| **(C6) Stay at home requirements** |  |  |  |  |
| 0 – no measures | 31.31 | 31.58 | 30.15 | 37.47 |
| 1 – recommend not leaving house | 26.60 | 27.34 | 25.89 | 24.11 |
| 2 – require not leaving house with exceptions for daily exercise, grocery shopping, and 'essential' trips | 35.34 | 37.70 | 40.20 | 34.22 |
| 3 – require not leaving house with minimal exceptions | 6.75 | 3.34 | 3.48 | 4.11 |
| **(C7) Restrictions on internal movement between cities/regions** |  |  |  |  |
| 0 – no measures | 34.01 | 47.17 | 50.63 | 49.40 |
| 1 – recommend not to travel between regions/cities | 17.10 | 17.61 | 17.02 | 15.48 |
| 2 – internal movement restrictions in place | 48.89 | 35.21 | 32.05 | 35.02 |
| **(C8) Restrictions on international travel for foreign travelers** |  |  |  |  |
| 0 – no restrictions | 5.71 | 0.30 | 0.44 | 9.35 |
| 1 – screening arrivals | 4.97 | 22.34 | 25.00 | 16.86 |
| 2 – quarantine arrivals from some or all regions | 11.26 | 27.97 | 27.06 | 20.06 |
| 3 – ban arrivals from some regions | 28.52 | 33.05 | 32.01 | 29.23 |
| 4 – ban on all regions or total border closure | 49.53 | 16.32 | 15.35 | 24.46 |
| **(H1) Presence of public information campaigns** |  |  |  |  |
| 0 – no COVID-19 public information campaign | 3.03 | 0.30 | 0.93 | 8.23 |
| 1 – public officials urging caution about COVID-19 | 5.32 | 5.89 | 6.31 | 6.37 |
| 2 – Coordinated public information campaigns | 91.65 | 93.79 | 92.35 | 85.26 |

Details of the statistical model-fitting methods we used are in the technical appendix. In summary, We scaled the Oxford Stringency Index to the range [0, 1] for the use in fractional regression modeling.[26] We estimated marginal effects associated with 1 new case per 100,000 people, 1 new death per 100,000 people, 1% larger GDP per capita, and 1% additional share of GDP spent on health. We report marginal effects instead of regression coefficients to enable more meaningful interpretations of the predicted change in the stringency score.

Our models used temporal lags to understand how rapidly governments enacted policies following new cases and deaths We fit separate models for the marginal effects based on 1- and 4-week lags between reporting a new case or death per 100,000 people and the associated average change in the stringency. To examine regional differences, we conducted separate analyses for each of the six WHO regions: Africa, Americas, Eastern Mediterranean, Europe, South-East Asia, and Western Pacific. A *p*-value less than 0.05 was considered statistically significant in all models.

## RESULTS

Policy stringency waned over time. For instance, all levels of schools and universities were required to close on 61.2% of the country-days in March–August 2020 compared to 20.8% in March–August 2021. However, some policies remained relatively unchanged. Restrictions on gatherings between 11 and 100 people were in place for 28.3% of the country-days in March–August 2020 compared to 31.7% in March– August 2021. The global median stringency score was higher for March–August 2020 (68.5) compared to September 2020–February 2021 (56.4) and March–August 2021 (54.6) (Table 1). The unadjusted median stringency score increased as new cases and deaths per 100,000 people increased (Figure 1).

**Figure 1.**
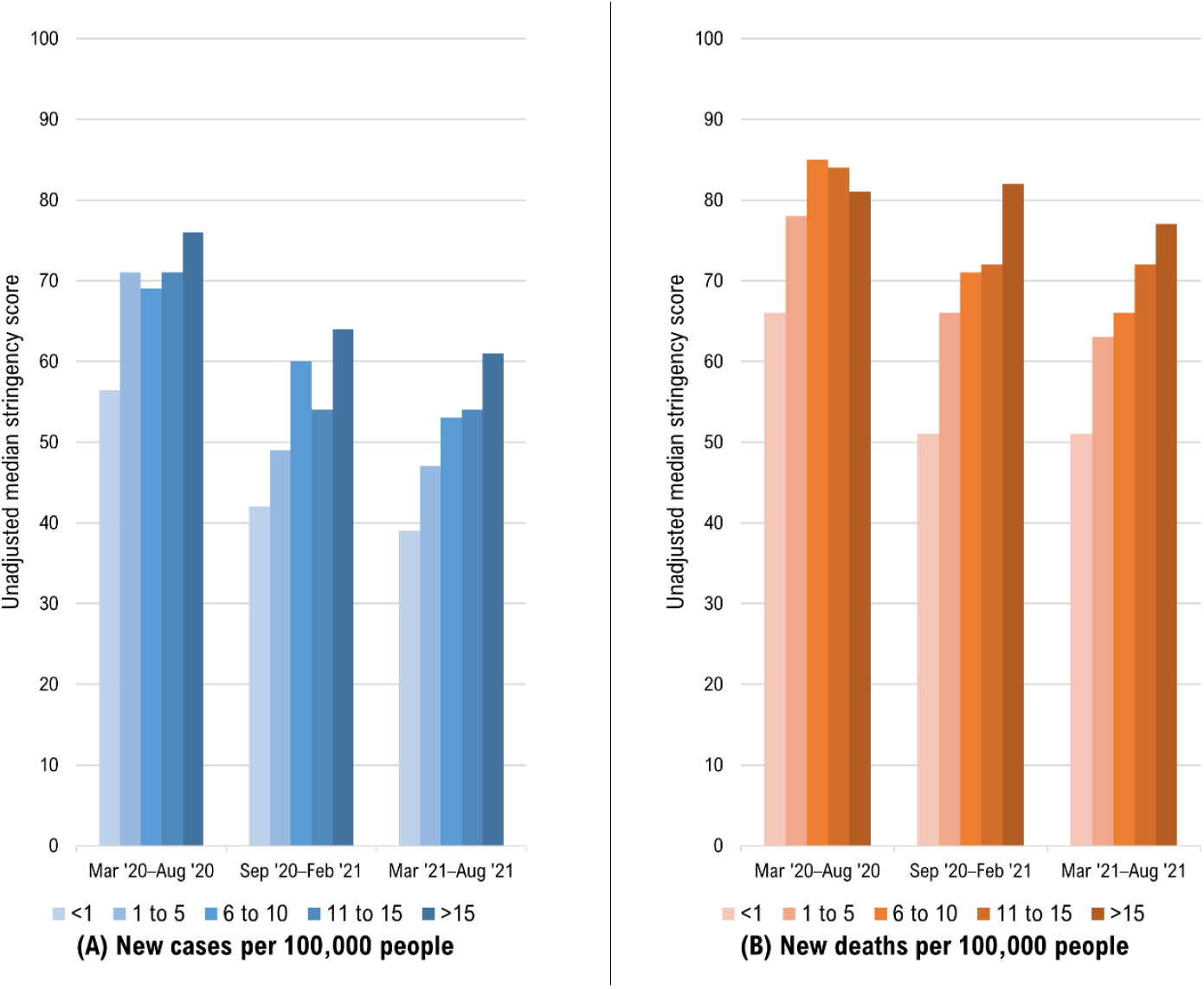
Changes in overall COVID-19 policy stringency as a function of new cases and deaths per 100,000 people in 175 countries and territories, March 2020—August 2021.

### Global findings

Within 1 week on average, 1 new case per 100,000 people was associated with a stringency increase of 0.11 units (95% Confidence Interval [CI]: 0.08 to 0.13) in March–August 2020, 0.02 units (0.01 to 0.02) in September 2020–February 2021, and 0.02 units (0.01 to 0.03) in March–August 2021. In the early period of the pandemic (March–August 2020), increased new cases were associated with more rapidly enacted stringent policies. (Table 2)

**Table 2.** Marginal effects of correlates on COVID-19 policy stringency in 175 countries and territories, March 2020—August 2021.

| Correlates | Global<br>Mar 2020 - Aug 2020 |  |  | Global<br>Sep 2020 - Feb 2021 |  |  | Global<br>Mar 2021 - Aug 2021 |  |  |
| --- | --- | --- | --- | --- | --- | --- | --- | --- | --- |
|  | ME | 95%CI |  | ME | 95%CI |  | ME | 95%CI |  |
| 1-week lag between reporting of new cases/deaths and policy stringency |  |  |  |  |  |  |  |  |  |
| New cases<sup>‡</sup> | 0.11 | 0.08 | 0.13 | 0.02 | 0.01 | 0.02 | 0.02 | 0.01 | 0.03 |
| New deaths<sup>‡</sup> | 2.09 | 1.33 | 2.85 | 1.30 | 0.96 | 1.64 | 0.71 | 0.36 | 1.06 |
| GDP per capita<sup>†</sup> | -0.02 | -0.03 | -0.02 | 0.03 | 0.02 | 0.03 | 0.02 | 0.02 | 0.02 |
| Health expenditure<sup>†</sup> | -1.21 | -1.51 | -0.90 | -0.92 | -1.17 | -0.68 | -0.89 | -1.11 | -0.68 |
| 4-week lag between reporting of new cases/deaths and policy stringency |  |  |  |  |  |  |  |  |  |
| New cases<sup>‡</sup> | 0.05 | 0.03 | 0.07 | 0.02 | 0.01 | 0.02 | 0.02 | 0.02 | 0.03 |
| New deaths<sup>‡</sup> | 0.84 | 0.45 | 1.23 | 1.09 | 0.72 | 1.46 | 0.24 | 0.00 | 0.47 |
| GDP per capita<sup>†</sup> | -0.02 | -0.02 | -0.01 | 0.03 | 0.03 | 0.04 | 0.02 | 0.01 | 0.02 |
| Health expenditure<sup>†</sup> | -1.00 | -1.26 | -0.74 | -0.83 | -1.09 | -0.57 | -0.92 | -1.14 | -0.69 |

*Note.* All findings are statistically significant, $p < .05$ . Green shading shows positive marginal effects, and yellow shading shows negative marginal effects. ME = marginal effects from three separate fractional regression models for each 6-month period; GDP = gross domestic product; CI = confidence interval
\*Epi-week 10 to 35 in 2020 used for March 2020—August 2020; epi-week 36 in 2020 to epi-week 8 in 2021 used for September 2020—February 2021; epi-week 9 in 2021 to epi-week 34 in 2021 used for March 2021—August 2021
<sup>‡</sup>1 new case per 100,000 population; 1 new death per 100,000 population
<sup>†</sup>1% increase in GDP per capita; 1% additional share of GDP on health expenditure

Within 1 week on average, 1 new death per 100,000 people was associated with a stringency increase of 2.1 units (1.3 to 2.9) in March–August 2020, 1.3 units (0.9 to 1.6) in September 2020–February 2021, and 0.7 units (0.4 to 1) in March–August 2021. Like new cases, in the early period of the pandemic, increased new deaths were associated with more rapidly enacted stringent policies (Table 2).

Although 1% larger GDP per capita was associated with less stringency in March–August 2020, it was associated with more stringency in the subsequent periods. A 1% additional share of GDP spent on health was associated with less stringency across all three periods (Table 2).

### Regional findings

We show the regional-specific marginal effects on policy stringency for a 1-week lag (Table 3) versus a 4-week lag (Table 4) after reporting new cases and deaths. An average increase of 1 new case per 100,000 people was associated with more stringent policies within 1 week in the Americas, Europe, and Western Pacific across all three periods. There was a slow policy response to new cases in Africa early in the pandemic. However, the association between new deaths and stringency was strongest in Africa early in the pandemic. Eastern Mediterranean was the only region where new cases were ever associated with less stringency (March 2021–August 2021).

**Table 3.**
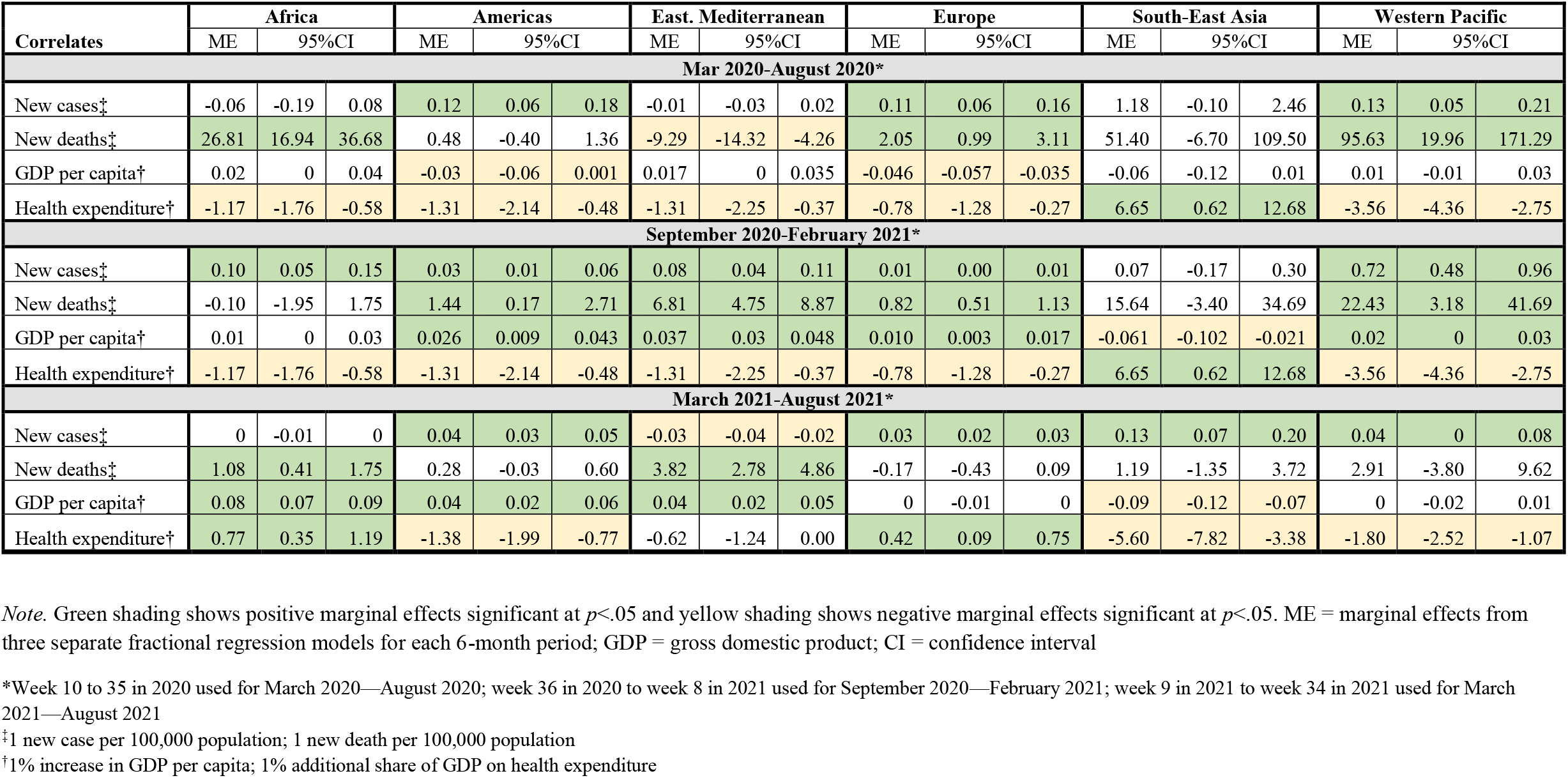
Regional marginal effects of correlates on COVID-19 policy stringency in 175 countries and territories, based on a 1-week lag after the reporting of cases and deaths, March 2020—August 2021.

**Table 4.** Regional marginal effects of correlates on COVID-19 policy stringency in 175 countries and territories, based on a 4-week lag after the reporting of cases and deaths, March 2020—August 2021.

|  | Africa |  |  | Americas |  |  | East. Mediterranean |  |  | Europe |  |  | South East Asia |  |  | Western Pacific |  |  |
| --- | --- | --- | --- | --- | --- | --- | --- | --- | --- | --- | --- | --- | --- | --- | --- | --- | --- | --- |
|  | ME | 95%CI |  | ME | 95%CI |  | ME | 95%CI |  | ME | 95%CI |  | ME | 95%CI |  | ME | 95%CI |  |
| March 2020-August 2020* |  |  |  |  |  |  |  |  |  |  |  |  |  |  |  |  |  |  |
| New cases‡ | 0.15 | 0.00 | 0.31 | 0.10 | 0.04 | 0.16 | 0.01 | 0 | 0.03 | -0.02 | -0.05 | 0.01 | 0.53 | -0.33 | 1.39 | 0.04 | -0.03 | 0.10 |
| New deaths‡ | -2.02 | -10.79 | 6.75 | 0.39 | -0.23 | 1.01 | -5.71 | -9.88 | -1.55 | 1.40 | 0.89 | 1.92 | 128.52 | 52.76 | 204.28 | 19.13 | -33.13 | 71.38 |
| GDP per capita† | 0.02 | 0 | 0.03 | -0.03 | -0.06 | -0.01 | 0.01 | 0 | 0.02 | -0.04 | -0.05 | -0.03 | -0.05 | -0.083 | -0.01 | 0.02 | 0 | 0.034 |
| Health expenditure† | -0.91 | -1.37 | -0.46 | -0.89 | -1.60 | -0.18 | -0.86 | -1.60 | -0.12 | -0.66 | -1.08 | -0.24 | 5.75 | 2.35 | 9.15 | -3.53 | -4.13 | -2.94 |
| September 2020-February 2021* |  |  |  |  |  |  |  |  |  |  |  |  |  |  |  |  |  |  |
| New cases‡ | 0.16 | 0.11 | 0.21 | 0.04 | 0.03 | 0.06 | 0.08 | 0.04 | 0.11 | 0.01 | 0.003 | 0.01 | 0.03 | -0.17 | 0.23 | 0.93 | 0.44 | 1.41 |
| New deaths‡ | -4.14 | -6.08 | -2.21 | 1.00 | 0.20 | 1.80 | 7.17 | 5.00 | 9.35 | 0.43 | 0.09 | 0.76 | 19.28 | -1.44 | 40.00 | 14.88 | -7.42 | 37.18 |
| GDP per capita† | 0.02 | 0 | 0.03 | 0.028 | 0.010 | 0.047 | 0.030 | 0.02 | 0.042 | 0.01 | 0 | 0.02 | -0.05 | -0.10 | 0 | 0.018 | 0.004 | 0.032 |
| Health expenditure† | -1.10 | -1.57 | -0.63 | -1.84 | -2.46 | -1.22 | -1.51 | -2.54 | -0.49 | -0.11 | -0.48 | 0.27 | -8.10 | -10.76 | -5.45 | -1.10 | -1.79 | -0.40 |
| March 2021-August 2021* |  |  |  |  |  |  |  |  |  |  |  |  |  |  |  |  |  |  |
| New cases‡ | 0 | 0 | 0.01 | 0.03 | 0.02 | 0.04 | -0.02 | -0.03 | -0.01 | 0.02 | 0.01 | 0.03 | 0.11 | 0.05 | 0.17 | 0.05 | 0 | 0.10 |
| New deaths‡ | -0.86 | -1.82 | 0.11 | 0.35 | -0.002 | 0.71 | 2.95 | 1.65 | 4.25 | -0.51 | -0.77 | -0.24 | -3.00 | -6.14 | 0.15 | 3.46 | -5.13 | 12.05 |
| GDP per capita† | 0.08 | 0.07 | 0.10 | 0.04 | 0.02 | 0.06 | 0.03 | 0.02 | 0.05 | -0.005 | -0.01 | 0 | -0.10 | -0.13 | -0.07 | 0 | -0.02 | 0.02 |
| Health expenditure† | 0.79 | 0.36 | 1.22 | -1.38 | -2.04 | -0.72 | -0.50 | -1.14 | 0.14 | 0.40 | 0.05 | 0.74 | -3.31 | -6.13 | -0.49 | -1.86 | -2.65 | -1.08 |

*Note.* Green shading shows positive marginal effects significant at $p < .05$ , and yellow shading shows negative marginal effects significant at $p < .05$ . ME = marginal effects from three separate fractional regression models for each 6-month period; GDP = gross domestic product; CI = confidence interval
ME = marginal effects from separate fractional regression models for each region; GDP = gross domestic product; CI = confidence interval
\*Week 10 to 35 in 2020 used for March 2020—August 2020; week 36 in 2020 to week 8 in 2021 used for September 2020—February 2021; week 9 in 2021 to week 34 in 2021 used for March 2021—August 2021
‡1 new case per 100,000 population; 1 new death per 100,000 population
†1% increase in GDP per capita; 1% additional share of GDP on health expenditure

GDP per capita did not have consistent associations with stringency across all periods within any region, and the pattern of associations was not the same for any two regions. Higher health expenditure was associated with less stringent policies within each region in March 2020–August 2020 and September 2020–February 2021, except for South-East Asia. Higher health expenditure was associated with less stringent policies within each region in March 2021–August 2021, except for Europe and the Eastern Mediterranean.

## DISCUSSION

To our knowledge, this is the first study to show the epidemiological drivers of COVID-19 policy stringency globally and within regions, adjusting for GDP per capita and health expenditure. We have empirically demonstrated that countries and territories responded more rapidly and strongly to the rise in new cases and deaths early in the COVID-19 pandemic than in subsequent periods. New deaths were more strongly associated with stringent policies than new cases across all periods examined. Despite African countries having weaker mortality surveillance systems [27], the association between new deaths and policy stringency was more potent in Africa early in the pandemic than other regions (except for Western Pacific). Health expenditure was negatively associated with policy stringency, suggesting that countries with less health expenditures may be more inclined to enact more stringent policies. GDP per capita did not have consistent patterns of associations globally or within regions.

Prior assessments have focused on understanding the effects of public health policies on COVID-19 epidemiological and clinical outcomes. The current evidence shows that public health policies are essential for controlling SARS-COV-2 transmission and will likely remain vital despite the increased accessibility of safe and effective COVID-19 vaccines.[7] In our study, we found that the overall stringency of policies decreased globally, coinciding with an increasing vaccine coverage worldwide, especially in high- and (upper-) middle-income countries. As an example, COVID-19 cases surged in Seychelles after the sudden relaxation of public health policies for non-pharmaceutical interventions in April 2021 before the detection of Omicron, despite rapidly scaling up vaccination coverage to 60%.[28] It has also been documented that maintaining stringent policies for prolonged periods also comes at a cost. School closures showed associations with adverse psychosocial outcomes among children during the pandemic.[29] More broadly, the COVID-19 pandemic has contributed to adverse mental health,[30] food insecurity,[31] and decreased utilization of primary healthcare services [32].

A critical gap in the current literature is the lack of an empirical understanding of how the reported new COVID-19 cases and deaths may influence the stringency of policies. Our study provides new evidence in the global landscape and within geographic regions. An empirical understanding of policy drivers can help public health officials anticipate policy responses in the COVID-19 pandemic and future health emergencies. Thus far, mathematical modelers have not calibrated their models to account for governments’ dynamic drivers of policy responses. Our results can help improve the parametrization of mathematical models attempting to forecast the course of a health emergency.

A prior study showed that countries with weaker health systems, such as those in low- and middle-income countries, were quicker to implement more stringent policies early in the pandemic.[3] High-income countries with more significant investments in their healthcare systems were better equipped to absorb the initial shocks of the COVID-19 burden. Globally, however, healthcare systems have been overwhelmed at peak times of hospitalizations during each COVID-19 wave.[33] We found that places with less health expenditure were more inclined to enact stringent policies. Countries with higher health expenditure may rely more on their capacity to provide medical care to patients instead of instituting stringent policies. Moreover, additional research is needed to understand to what extent countries with weak mortality surveillance may have opted for less stringent policies because of their under-detection of COVID-19 deaths. Unpredictable political and economic considerations make it difficult to anticipate how governments react to this pandemic’s policymaking decisions or future health emergencies.

Our study has several limitations. Firstly, we cannot discern the enforcement of or adherence to government policies. In addition, we did not account for the potential influence of geographic proximity between countries. Although hospitalizations and hospital bed capacities likely mediated policy stringency, standardized global data on these possible mediating variables are lacking and therefore not included in our models.

## CONCLUSION

The findings from this study reinforce the need for enhanced mortality surveillance to ensure policy alignment during health emergencies. Timely detection of excess deaths may help policymakers communicate their policy decisions more effectively to the public. Countries with lesser health expenditure may be more inclined to enact stringent policies during health emergencies than countries with more significant health expenditure as a percent of GDP. Our findings call for public health authorities to tailor their support to policymakers to ensure alignment between the stringency of containment policies and the public health requirements of the health emergency.

## Data Availability

All data produced in the present study are available upon reasonable request to the authors.

## Acknowledgements

We acknowledge various technical support from Dr. Barbara Marston from the U.S. Centers for Disease Control and Prevention and Dr. Oliver Morgan and Dr. Olivier Le Polain from the World Health Organization.

## Contributors

MFJ, ZZ, SAN, DP, AAN, MW, and HN conceptualized the study. MFJ, ZZ, HN, DP, AH, SBO, NTB made methodological contributions. MFJ conducted the literature review with additional literature identified by ZZ, AH, HN, LS, and WG. ZZ and MFJ analyzed, curated, and visualized the data with substantial contributions from MW, HN, AAN, and LS. MFJ, ZZ, SAN, DP, AAN, AH, MW, LS, WG, SBO, NTB, and HN contributed to the interpretation of the results. MFJ and ZZ drafted the initial manuscript with contributions from SAN, DP, AAN, AH, MW, LS, WG, SBO, NTB, and HN. All authors contributed to reviewing and editing the manuscript. The final manuscript was read and approved by all authors.

## SUPPLEMENTARY MATERIAL

We fit several fixed-effects fractional regression models using a quasi-likelihood method with a logit link to examine associations between weekly new cases/deaths and policy stringency, adjusting for GDP per capita and health expenditure in 175 countries and territories. Our fractional regression models accounted for a country’s/territory’s specific and time-varying policy stringency on a nonlinear-trend basis by including interaction terms for (1) country × time and (2) country × time-squared. The time-varying effect is measured weekly over 6-month periods. The time is mean-centered for the avoidance of multicollinearity between time and time-squared. The mean centering is done by subtracting the middle week number of the studied period from each week number. The country-specific intercept in the fixed-effects model, was replaced by other time-invariant GDP per capita and health expenditure on country level to avoid the dummy trap problem (perfect multicollinearity).

The stringency index, scaled in the interval [0,1], for country *i* at time *t* was modeled as

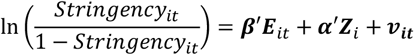

where ***E***_*it*_ is the vector of weekly COVID-19 cases and deaths, ***Z***_*i*_ the vector of observed and time-invariant country-specific GDP per capita and health expenditure, and ***β*** and ***α*** are unknown regression parameters to be estimated. Also, ***ν***_***it***_ is the unobservable time-varying country’s specific performance at time t, modeled as:

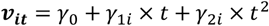

with unknown parameters *γ*_0_, *γ*_1*i*_ and *γ*_2*i*_, to be estimated for country *i*. Alternatively, the above country-specific performance at time t is presented as in the fixed-effects notation. It can be equivalently represented as:

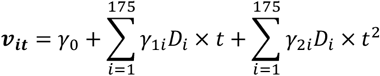

where *D*_*i*_ is the ith country dummy variable.

For sensitivity analysis, the time-invariant predictors are used in a random-effects beta regression model, using glmmTMB library in R statistical software package version 1.1.2.3 (R Foundation for Statistical Computing, Vienna, Austria). The beta regression model was fitted as: 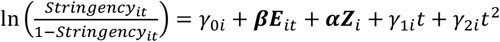, with *γ*_*ji*_ = *γ*_*j*_ + *u*_*ji*_, *j* = 0,1,2, and *u*_*ji*_ a random variable.

The results of the beta regression and the fractional regression were consistent whenever the beta regression is applicable. The beta regression was not appropriate for extreme situations with 0 or 100 stringency scores (0,1 on a fractional scale). Such extreme situations were rare in the dataset, especially after March—August 2020 period. We only report here the results from the fractional regression.

## REFERENCES

1 Desson Z, Weller E, McMeekin P, Ammi M. An analysis of the policy responses to the COVID-19 pandemic in France, Belgium, and Canada. Health Policy Technol. 2020;9:430–46.

2 Chen S, Guo L, Alghaith T, Dong D, Alluhidan M, Hamza MM, et al. Effective COVID-19 Control: A Comparative Analysis of the Stringency and Timeliness of Government Responses in Asia. Int J Environ Res Public Health. 2021;18.

3 Walker PGT, Whittaker C, Watson OJ, Baguelin M, Winskill P, Hamlet A, et al. The impact of COVID-19 and strategies for mitigation and suppression in low-and middle-income countries. Science. 2020;369:413–22.

4 Kissler SM, Tedijanto C, Goldstein E, Grad YH, Lipsitch M. Projecting the transmission dynamics of SARS-CoV-2 through the postpandemic period. Science. 2020;368:860–8.

5 Fuller JA, Hakim A, Victory KR, Date K, Lynch M, Dahl B, et al. Mitigation Policies and COVID-19-Associated Mortality - 37 European Countries, January 23-June 30, 2020. MMWR Morb Mortal Wkly Rep. 2021;70:58–62.

6 World Health Organization. Tracking Public Health and Social Measures: Global Databaset. 2020. Available: https://www.who.int/emergencies/diseases/novel-coronavirus-2019/phsm.

7 Rella SA, Kulikova YA, Dermitzakis ET, Kondrashov FA. Rates of SARS-CoV-2 transmission and vaccination impact the fate of vaccine-resistant strains. Sci Rep. 2021;11:15729.

8 World Health Organization. Update on Omicron. 2021. Available: https://www.who.int/news/item/28-11-2021-update-on-omicron.

9 World Health Organization. Vaccine equity. 2021. Available: https://www.who.int/campaigns/vaccine-equity.

10 World Health Organization. Coronavirus disease (COVID-2019) situation reports. 2020. Available: https://www.who.int/emergencies/diseases/novel-coronavirus-2019/situation-reports.

11 Chen CWS, Lee S, Dong MC, Taniguchi M. What factors drive the satisfaction of citizens with governments’ responses to COVID-19? International journal of infectious diseases : IJID : official publication of the International Society for Infectious Diseases. 2021;102:327–31.

12 Rostami A, Sepidarkish M, Leeflang MMG, Riahi SM, Nourollahpour Shiadeh M, Esfandyari S, et al. SARS-CoV-2 seroprevalence worldwide: a systematic review and meta-analysis. Clin Microbiol Infect. 2021;27:331–40.

13 Bobrovitz N, Arora RK, Cao C, Boucher E, Liu M, Donnici C, et al. Global seroprevalence of SARS-CoV-2 antibodies: A systematic review and meta-analysis. PLoS One. 2021;16:e0252617.

14 Lau H, Khosrawipour T, Kocbach P, Ichii H, Bania J, Khosrawipour V. Evaluating the massive underreporting and undertesting of COVID-19 cases in multiple global epicenters. Pulmonology. 2021;27:110–5.

15 Devex. Data around COVID-19 is a mess and here’s why that matters. 2020. Available: https://www.devex.com/news/data-around-covid-19-is-a-mess-and-here-s-why-that-matters-97077.

16 TheEconomist. Covid-19 presents economic policymakers with a new sort of threat: It mixes demand and supply effects. 2020. Available: https://www.economist.com/finance-and-economics/2020/02/20/covid-19-presents-economic-policymakers-with-a-new-sort-of-threat.

17 Friedler A. Sociocultural, behavioural and political factors shaping the COVID-19 pandemic: the need for a biocultural approach to understanding pandemics and (re)emerging pathogens. Global public health. 2021;16:17–35.

18 Papke LE, Wooldridge JM. Econometric methods for fractional response variables with an application to 401(k) plan participation rates. Journal of Applied Econometrics. 1996;11:619–32.

19 Cross M, Ng SK, Scuffham P. Trading Health for Wealth: The Effect of COVID-19 Response Stringency. Int J Environ Res Public Health. 2020;17.

20 cesinfo. Health expenditures and the effectiveness of COVID-19 prevention in international comparison. 2021. Available: https://www.cesifo.org/en/publikationen/2021/working-paper/health-expenditures-and-effectiveness-covid-19-prevention.

21 UnitedNations. Expidture on health. 2020. Available: http://data.un.org/_Docs/SYB/CSV/SYB63_325_202009_Expenditure%20on%20Health.csv.

22 Oxford. Coronavirus government response tracker. 2020. Available: https://www.bsg.ox.ac.uk/research/research-projects/coronavirus-government-response-tracker.

23 Hale T, Angrist N, Goldszmidt R, Kira B, Petherick A, Phillips T, et al. A global panel database of pandemic policies (Oxford COVID-19 Government Response Tracker). Nature human behaviour. 2021;5:529–38.

24 World Health Organization. COVID-19 Global Data. 2021. Available: https://covid19.who.int/WHO-COVID-19-global-data.csv.

25 UnitedNations. Gross domestic product per capita. 2020. Available: http://data.un.org/_Docs/SYB/CSV/SYB63_230_202009_GDP%20and%20GDP%20Per%20Capita.csv.

26 Mullahy J. Multivariate fractional regression estimation of econometric share models. J Econom Method. 2015;4:71–100.

27 Africa CDC. Mortality Surveillance Programme. 2018. Available: https://africacdc.org/programme/surveillance-disease-intelligence/mortality-surveillance-programme/.

28 Vilches TN, Sah P, Abdollahi E, Moghadas SM, Galvani AP. Importance of non-pharmaceutical interventions in the COVID-19 vaccination era: A case study of the Seychelles. Journal of global health. 2021;11:03104.

29 Singh S, Roy D, Sinha K, Parveen S, Sharma G, Joshi G. Impact of COVID-19 and lockdown on mental health of children and adolescents: A narrative review with recommendations. Psychiatry Res. 2020;293:113429.

30 World Health Organization. Mental Health and COVID-19: Early evidence of the pandemic’s impact. 2022. Available: https://www.who.int/publications/i/item/WHO-2019-nCoV-Sci_Brief-Mental_health-2022.1.

31 Picchioni F, Goulao LF, Roberfroid D. The impact of COVID-19 on diet quality, food security and nutrition in low and middle income countries: A systematic review of the evidence. Clin Nutr. 2021.

32 Aranda Z, Binde T, Tashman K, Tadikonda A, Mawindo B, Maweu D, et al. Disruptions in maternal health service use during the COVID-19 pandemic in 2020: experiences from 37 health facilities in low-income and middle-income countries. BMJ Glob Health. 2022;7.

33 Miller IF, Becker AD, Grenfell BT, Metcalf CJE. Disease and healthcare burden of COVID-19 in the United States. Nat Med. 2020;26:1212–7.

